# Exploratory Randomised Trial of Tranexamic Acid to Decrease Postoperative Delirium in Adults Undergoing Lumbar Fusion: A trial stopped early

**DOI:** 10.1101/2024.10.16.24315638

**Authors:** Bradley J. Hindman, Catherine R. Olinger, Royce W. Woodroffe, Mario Zanaty, Carolina Deifelt Streese, Zeb R. Zacharias, Jon C. D. Houtman, Linder H. Wendt, Patrick P. Ten Eyck, Debra J. O’Connell-Moore, Emanuel J. Ray, Sarah J. Lee, Daniel F. Waldschmidt, Lauren G. Havertape, Lanchi B. Nguyen, Pei-fu Chen, Matthew I. Banks, Robert D. Sanders, Matthew A. Howard

**Author notes:** Corresponding author: Bradley J. Hindman. Duke University School of Medicine, Durham, North Carolina, USA. California Health Sciences University College of Osteopathic Medicine, Clovis, California, USA. The University of Iowa Roy J. and Lucille A. Carver College of Medicine, Iowa City, Iowa, USA.

## Abstract

**Background:** Postoperative delirium may be mediated by perioperative systemic- and neuro-inflammation. By inhibiting the pro-inflammatory actions of plasmin, tranexamic acid (TXA) may decrease postoperative delirium. To explore this hypothesis, we modified an ongoing randomised trial of TXA, adding measures of postoperative delirium, cognitive function, systemic cytokines, and astrocyte activation.

**Methods:** Adults undergoing elective posterior lumbar fusion randomly received intraoperative intravenous TXA (n=43: 10 mg kg^-1^ loading dose, 2 mg kg^-1^ h^-1^ infusion) or Placebo (n=40). Blood was collected pre- and at 24 h post-operatively (n=32) for biomarkers of systemic inflammation (cytokines) and astrocyte activation (S100B). Participants had twice daily delirium assessments using the 3-minute diagnostic interview for Confusion Assessment Method (n=65). Participants underwent 4 measures of cognitive function preoperatively and during post-discharge follow-up.

**Results:** Delirium incidence in the TXA group (7/32=22%) was not significantly less than in the Placebo group (11/33=33%); *P*=0.408, absolute difference=11%, relative difference=33%, effect size = −0.258 (95% CI −0.744 to 0.229). In the Placebo group (n=16), delirium severity was associated with the number of instrumented vertebral levels (*P*=0.001) and with postoperative interleukin −8 and −10 concentrations (*P*=0.00008 and *P*=0.005, respectively) and these associations were not significantly modified by TXA. In the Placebo group, delirium severity was associated with S100B concentration (*P*=0.0009) and the strength of the association was decreased by TXA (*P*=0.002).

**Conclusions:** A potential 33% relative decrease in postoperative delirium incidence justifies an adequately powered clinical trial to determine if intraoperative TXA decreases delirium in adults undergoing lumbar fusion.

## Introduction

Postoperative delirium occurs in 15-40% of patients, varying with patient characteristics and type of procedure.^1,2^ Postoperative delirium is associated with a greater incidence of pre-discharge adverse events,^1,3^ greater length of hospital stay,^1,3^ greater incidence of non-home discharge,^1,3^ greater readmissions,^1,3^ greater short- and long-term costs,^4^ greater mortality,^1^ and long-term cognitive decline.^5,6^

Postoperative delirium has been intensively studied in numerous preclinical models and clinical studies. These studies indicate that many factors contribute to postoperative delirium.^1,7,8^ Among these factors is the brain’s response to systemic injury. Tissue injury— occurring with surgical procedures, trauma, or burns—results in inflammation at the site of injury as well as increased circulating concentrations of pro-inflammatory mediators.^9^ These circulating mediators trigger inflammatory responses within the brain (neuroinflammation),^10,11^ resulting in changes in behaviour and cognition. In principle, decreasing the magnitude of systemic inflammatory response and/or decreasing subsequent neuroinflammatory response should decrease the incidence and/or severity of postoperative delirium; this has been demonstrated in animal models.^12,13^

Tranexamic acid (TXA) is a lysine analog that competitively blocks plasminogen binding to fibrin, decreasing or preventing fibrinolysis.^14,15^ In common clinical use for decades, TXA is often administered off-label to decrease perioperative blood loss.^16–18^ It is now known that many cell types have plasmin(ogen) receptors,^19^ and that plasmin(ogen) has many physiologic roles in addition to fibrinolysis,^20,21^ including modulating neuroinflammation.^22^ By inhibiting lysine-dependent plasmin(ogen) binding to its receptor, TXA has the potential to decrease plasmin(ogen)-mediated inflammatory responses.^21,23^ For example, in some clinical studies, TXA decreases postoperative proinflammatory cytokine concentrations.^24^ Plasmin also plays a role in modulating the endothelial blood-brain barrier^25^ and animal studies suggest plasmin inhibition may prevent blood-brain barrier injury.^26^ Because of its established capacity to decrease blood loss and transfusion^27^ and its potential to decrease plasmin-mediated inflammation and blood-brain barrier injury,^28^ some authors have hypothesized that TXA might decrease postoperative delirium.

To explore this hypothesis, we modified an ongoing randomised trial of TXA in adults undergoing lumbar fusion to include additional outcome measures, specifically, postoperative delirium, biomarkers of systemic- and neuro-inflammation, and post-discharge cognitive function.

## Methods

### Participant eligibility, original protocol, and outcomes

This study was registered with ClinicalTrials.gov prior to patient enrollment (NCT04272606; February 17, 2020) and was approved by the University of Iowa Institutional Review Board (IRB) for Human Subjects (#201912099; April 23, 2020). All participants provided written informed consent. Enrollment was delayed because of the onset of the COVID pandemic; the first participant enrolled on November 5, 2020. The original primary outcome measures were postoperative blood loss and transfusion. The original projected enrollment of 150 participants was powered to detect a 50% decrease in mean intraoperative blood loss in participants who received TXA based on a historical control value (mean±SD) of 600±400 mL (alpha = 0.05, 90% power).

Participants were adults (age 18 to 90 years) undergoing elective open posterior lumbar, thoracolumbar, or lumbosacral fusion to treat symptoms of degenerative spine disease at the University of Iowa. Participant’s sex was as designated in their medical record. There were 29 exclusion criteria that were intended to decrease risks of potential TXA side effects including 1) prior seizures; 2) preoperative creatinine >133 μmol L^-1^; 3) any prior intracranial or ophthalmologic vascular event; 4) any prior deep venous thrombosis or other condition related to hypercoagulability; 5) preoperative hormonal therapy (*e.g.*, oral contraceptives or testosterone); 6) abnormal color vision; and 7) any intravascular stents. Other exclusion criteria were intended to decrease the risk of large surgical blood loss and/or transfusion including: 1) preoperative haemoglobin <80 g L^-1^ or platelet count <1.5 x 10^5^ per μL; or 2) prothrombin time > 15 seconds or activated partial thromboplastic time >38 seconds. Potential participants who had severe cardiac, respiratory, or hepatic disorders were also excluded. Exclusion in the interval between consent and the procedure occurred when a new or previously unrecognized exclusion criterion was discovered.

Participants were randomised by the Investigational Pharmacy to one of two study medication groups, either TXA or Placebo, using a block size of 10. Study medications were prepared by an investigational pharmacist and provided in bags labeled as study medication. All participants, clinical care providers, and research personnel were blinded to the treatment assignments until all outcome determinations for all participants for the entire study were completed. TXA was given intravenously as a loading dose (10 mg kg^-1^) started approximately 20 minutes before incision, followed by continuous intraoperative infusion (2 mg kg^-1^ h^-1^) which was discontinued when wound closure was complete.^29^ Participants randomised to Placebo received equivalent volumes of saline.

Participants received routine (non-standardized) postoperative care determined by their surgical teams. Participants were evaluated daily by study personnel for the first five postoperative days or until discharge, whichever came first. In addition to postoperative delirium assessments (see Delirium Assessments), predefined safety outcome measures included: 1) postoperative day 1 creatinine increase >20% from the preoperative value; 2) any thromboembolic event (*e.g.*, deep venous thrombosis or pulmonary embolism); 3) any surgical wound abnormality (*e.g.*, wound healing problem or infection); 4) any visual symptoms; and/or 5) seizures. Participants had routine post-discharge follow-up surgical clinic visits at approximately 6 weeks and 3 months after the procedure and were evaluated by study personnel during these visits.

### Protocol and outcomes modifications

With prior IRB approval, a series of study modifications were made. On July 26, 2021 postoperative delirium was added as a secondary outcome measure and preoperative and post-discharge cognitive testing was added. Delirium was added as an outcome measure based on the report by Taylor and colleagues in which the anti-inflammatory properties of TXA were proposed to potentially decrease delirium incidence.^28^ On August 24, 2021 a prior eligibility requirement for fusion to include ≥2 interspaces was removed and projected enrollment was increased from 150 to 300 participants. The increase in enrollment was made because it was anticipated that inclusion of participants undergoing less extensive procedures would decrease blood loss in both groups and, consequently, increase the number of participants needed to detect TXA’s potential effect on blood loss and transfusion. On November 2, 2021 pre- and post-operative blood collection was added in order to measure biomarkers of systemic inflammation (plasma cytokines, T-lymphocyte immunophenotypes) and astrocyte activation (S100B protein). On March 25, 2022 delirium was changed from a secondary to a primary outcome measure, and biomarkers and cognitive performance were added as secondary outcome measures.

### Blood loss and red blood cell transfusion

Intraoperative estimated blood loss was obtained from the anaesthesia record. In 4 participants for whom blood loss was not recorded, the corresponding surgeon’s procedure note was reviewed and, in all cases, the note reported intraoperative blood loss was “none/minimal.” In these 4 participants (2 from each group), a default intraoperative blood loss of zero (0) mL was used for analysis.

Postoperative blood loss was based on wound drain output. With the exception of 4 participants (TXA [n = 1]; Placebo [n = 3]), subfascial wound drains (Hemovac^®^, Zimmer Biomet) were inserted during surgical closure. Drain collection chambers were emptied and volumes were recorded by nursing staff every 8 hours. Drain output from the end of the procedure to 06:59 AM the next day (postoperative day 1) was designated as wound output (blood loss) for postoperative day 0. Drain output for postoperative day 1 began at 07:00 AM and consisted of the sum of all volumes for the next 24 hours. Drains were removed when their output was <50 mL per 8-hour period. The duration of postoperative wound drain placement was defined as the last postoperative day during which the drain was present, even if the drain was not present for the entire 24-hour period. Postoperative blood loss equaled the total wound drain output obtained during postoperative days 0-3.

To adjust for the extent of intraoperative tissue injury, intra- and post-operative blood loss values were divided by the number of instrumented vertebral levels and is reported as mL per instrumented level. Intraoperative packed red blood cell transfusion was defined as occurring if one or more units were started in the operating room as documented on the anaesthesia record. Red blood cell transfusions started after participants left the operating room were designated as postoperative transfusions.

### Delirium assessments

Starting on postoperative day 1, participants were evaluated twice daily (at approximately 8-10 AM and 3-5 PM) by study personnel using the 20-item 3-minute diagnostic interview for Confusion Assessment Method (3D-CAM)^30,31^ as described in 3D-CAM Training Manuals 4.1 and 5.3.^32^ Delirium assessments were made through the afternoon of postoperative day five or the morning of the day of discharge, whichever came first. A diagnosis of delirium required the simultaneous presence of 3D-CAM feature 1 (acute onset or fluctuating course; at least 1 of 6 items) and feature 2 (inattention; at least 1 of 6 items) and either feature 3 (disorganized thinking; at least 1 of 6 items) or feature 4 (altered level of consciousness; at least 1 of 2 items). For each 3D-CAM exam, a delirium severity score was calculated *post hoc* as the sum of positive items (range 0 to 20 points).^33^

When participants were intubated or receiving sedation in an intensive care unit, the Confusion Assessment Method for the Intensive Care Unit (CAM-ICU) delirium assessment instrument was used.^34^ The CAM-ICU provides a delirium diagnosis but not a severity score. In instances of missing 3D-CAM or CAM-ICU evaluations, medical records were retrospective reviewed for the presence of Delirium Observation Screening Scale (DOSS) scores^35,36^ which were recorded every 12 hours as part of routine nursing practice in patients aged 65 years or older. Using a DOSS score ≥3 points (out of a maximum of 13) as the diagnostic threshold,^37^one participant in the Placebo group had a delirium diagnosis based on DOSS scores.

Participants were considered to have postoperative delirium if diagnostic criteria were satisfied on at least one evaluation prior to discharge. Delirium day of onset was the first postoperative day in which any delirium examination met the diagnostic criteria. The delirium onset severity score was the severity score for the first delirium diagnosis. The number of days with delirium was the sum of all days during which participants had at least one positive delirium exam; days of delirium were not required to be consecutive. The maximum delirium severity score was the maximum 3D-CAM severity score on any day, irrespective of whether the exam met the diagnostic criteria for delirium. Average delirium severity was the average of all delirium severity scores from all exams on all days, regardless of delirium diagnosis.

Among the 68 participants who had consented to postoperative delirium assessments (TXA [n = 34]; Placebo [n = 34]), 3 did not have any delirium assessments. Two participants (1 in each group) were discharged on the morning of postoperative day 1 before 3D-CAM exams were administered. A third participant (TXA group) withdrew their consent on postoperative day 1 prior to their delirium exam. One TXA participant withdrew their consent late on postoperative day 3; delirium data collected before their withdrawal was used in the analysis. Thus, delirium assessments were made in 65 participants (TXA [n = 32]; Placebo [n = 33]).

### Cognitive testing

Four cognitive tests were administered preoperatively and during post-discharge follow-up exams at approximately 6 weeks and 3 months after the procedure; see Results. The Telephone Interview of Cognitive Status—modified (TICSm) assesses global cognition, with an emphasis on learning and memory with scores ranging from 0 to 50 (best possible). TICSm is associated with both short- and long-term memory function^38^ and can distinguish among individuals who have normal cognition, mild cognitive impairment, or dementia.^39^

Trail Making Tests (TMT’s), Parts -A and -B, assess speed of cognitive processing and executive functioning. Part-A (TMT-A) primarily tests visual search and motor speed, whereas Part-B (TMT-B), which is more difficult, tests higher level (executive) abilities such as mental flexibility.^40,41^ Less time to complete a TMT indicates better performance. Raw performance scores were converted to *z*-scores using age- and education-matched population means and standard deviations.^42^ A *negative z*-score indicates the participant took less time to complete the test relative to their matched reference population (*i.e.*, better performance).

Controlled Oral Word Association (COWA) tests are verbal fluency tests measuring spontaneous production of words belonging to the same category (*e.g*., animals) or beginning with some designated letter; in this study, the letters C, F, and L. COWA tests language and/or executive function,^43^ and is impaired in setting of mild (preclinical) cognitive impairment.^43,44^ Participants had 1 minute to name as many words as possible beginning with the first letter. The procedure was then repeated for the remaining two letters. Raw performance scores were converted to *z*-scores using age- and education-matched population means and standard deviations.^42^ In contrast to the TMT’s, a *positive z*-score indicates a greater number of words relative to the matched reference population (*i.e.,* better performance).

Among the 68 participants who had consented to preoperative and post-discharge cognitive testing, 2 from the TXA group withdrew from the study on postoperative days 1 and 3 (noted above), such that post-discharge cognitive testing was not done. Among the remaining 66 participants, the following underwent cognitive testing both preoperatively and during post-discharge follow-up: TICSm (n = 55), TMT-A (n = 54), TMT-B (n = 51), and COWA (n = 37).

With small variations among the 4 tests, participants underwent post-discharge testing either once (∼60% of the participants) or twice (∼40% of the participants). When tested twice, the second post-discharge test was compared to the preoperative test. With small variations among the 4 tests, post-discharge cognitive testing took place 12 (7, 18) (median [25^th^, 75^th^ percentile]) weeks after the day of the procedure.

### Biomarkers of systemic inflammation and neuroinflammation

Whole blood for plasma and peripheral blood mononuclear cells (PBMC) was collected on the day of the procedure before incision (preoperative) and approximately 24 hours after the end of the procedure (postoperative). Blood was collected in Vacutainer® Cell Preparation Tubes with Sodium Heparin (CPT™). CPT tubes were centrifuged at room temperature in a horizontal rotor for a minimum of 15 minutes at 1500 g. The top layer (plasma) was pipetted into cryovials and stored at −80°C until analysis. The second layer (PBMCs) was pipetted into 15 mL conical tubes and washed 2 times with 1X Phosphate Buffered Saline (PBS). Additional Methods and Results regarding perioperative T-lymphocyte immunophenotypes and their relationships to TXA administration and delirium will be reported separately in a future publication.

All biomarker analyses were performed in the Human Immunology Core of the University of Iowa Holden Comprehensive Cancer Center. As a measure of systemic inflammation, plasma cytokine concentrations for interleukins (IL) −6, −8, and −10, and tumor necrosis factor, alpha (TNFα) were determined using a custom Milliplex Human Cytokine/Chemokine/Growth factor Assay (EMD Millipore, Burlington, MA) following the manufacturer’s instructions. As a measure of neuroinflammation (astrocyte activation), plasma was analyzed for S100B protein using the Human S100B DuoSet enzyme linked immunosorbent assay (R&D Systems, Minneapolis, MN) following the manufacturer’s instructions (The lower limits of detection of each assay are reported in Supplementary data Table S1). For the Milliplex, cytokine data were acquired using a BioRad Bio-Plex 200 and analyzed using BioRad Bio-Plex Manager Software (BioRad, Hercules, CA). For the S100B immunoassay, data were collected using a SpectraMax iD5 Multi-Mode Microplate Reader (Molecular Devices, San Jose, CA) and analyzed using GraphPad Prism software (GraphPad, Boston, MA).

Among the 43 participants who consented to perioperative blood sampling, blood for plasma biomarker analysis was collected preoperatively in 32 and postoperatively in 33. During statistical analysis of cytokine data, it was determined that one participant (TXA group) was an extreme upper outlier for all preoperative cytokine concentrations (data available but not shown). For this reason, this participant was excluded from all pre-*vs* post-operative biomarker analyses. Thus, there were a total of 32 participants with postoperative plasma samples (TXA [n=16]; Placebo [n=16]) and 31 participants who had paired pre- and post-operative plasma samples (TXA [n = 16]; Placebo [n = 15]). Among 31 preoperative plasma samples, IL-6 was undetectable in 2. For these 2 participants (1 in each group), a default preoperative IL-6 concentration of zero (0) pg mL^-1^ was used for analysis

### Nesting of delirium exploratory study within a trial that stopped early

Enrollment was stopped early (February 27, 2023; 123 enrolled) as the consequence of an *ad hoc* interim analysis of postoperative delirium incidence conducted on February 16, 2023. The interim analysis was performed because of the findings from a retrospective observational study from our group.^45^ In brief, the observational study included 570 patients who had undergone elective lumbar fusion, 169 (30%) of whom had received TXA. Without covariate adjustment, patients who had received TXA had an equivalent incidence of delirium when compared to those who did not, 20% *vs* 22%, respectively; *P*=0.60. However, with a propensity score-based analysis to adjust for variation in risk factors, patients who received TXA had a significantly lower incidence of delirium compared to those who did not, 14% *vs* 21%, respectively; *P*=0.004.^45^ We could not be sure which of these two observational results was correct—either TXA had *no effect* on delirium or TXA reduced delirium by 33%. A clinical trial to demonstrate TXA decreases delirium incidence from 21% (control) to 14% (alpha=0.05, power=80%) would require 462 participants per group; 924 in total. Thus, our observational data indicated our ongoing TXA trial was futile^46^ because either: 1) TXA has no effect on delirium; or 2) our trial was underpowered (with a planned enrollment of 300 participants instead of the required 924).

Therefore, to determine which was the case, an *ad hoc* interim analysis of this study was performed. Only the analysts (PT, WL) were made aware of individual participants’ treatment assignments. Interim results were reported only in aggregate form (TXA *vs* Placebo) without individual participant data. As reported in Results, the postoperative delirium incidence was not significantly less in the TXA group (7/32=22%) *vs* the Placebo group (11/33=33%); *P*=0.408. A clinical trial to demonstrate TXA decreases delirium incidence from 30% (control) to 20% (alpha = 0.05, power = 80%) would require 294 participants per group; 588 in total. Given our rates of enrollment and randomization, we concluded we could not randomize the required number of participants within a practical time frame. Consequently, we stopped the trial early. Reinforcing this decision was publication in January 2023 of a meta-analysis reporting the effectiveness of TXA to decrease blood loss in this specific patient population (posterior lumbar interbody fusion).^18^

### Statistical analysis

All analyses were conducted using R statistical software (version 4.3.1). Continuous variables were summarized using medians and interquartile ranges and comparisons between groups were made using Wilcoxon rank sum or Kruskal-Wallis tests. Pairwise analyses within a group were made using the Wilcoxon signed rank test. When comparing postoperative blood loss per instrumented level between treatment groups, Cohen’s d effect size along with its corresponding interval was also provided. Categorical variables are summarized using counts and percentages, and comparisons between groups are performed using Pearson χ^2^ test or Fisher exact test. Effect sizes and their corresponding confidence intervals were also provided when comparing delirium incidence between the treatment groups. Variables were also compared between groups by calculating standardized mean differences. Kendall’s tau (τ) was used as a non-parametric test of association between two variables.

As shown in Results, many outcome measures were associated with the number of instrumented vertebral levels (*e.g*., blood loss, delirium severity, postoperative cytokine concentrations). For this reason, in all regression models of associations between independent variables (*e.g*., group [TXA *vs* Placebo]) and outcomes (*e.g*., delirium severity), the number of instrumented levels was included as a second independent variable, and the interaction of the two independent variables was also included in the model.

*P* values are two-sided and the threshold for significance was <0.05 without adjustment for multiple comparisons. *P* values are reported to three decimal places unless < 0.001. P values less than 0.0000001 are reported as <0.0000001.

## Results

### Participant characteristics

Among 123 consenting participants, 37 (30%) were excluded prior to their procedure without being randomised (Fig 1). The most common reason for exclusion was detection of an exclusion criterion in the interval between consent and the day of their procedure (n=14), followed by procedure cancellation (n=8), participant withdrawal from the study (n=5), and the procedure changed to a minimally invasive procedure (n=3). Among 86 participants randomised to receive study medication, 3 did not receive it and were withdrawn from the study (no intra- or post-operative data were obtained): 2 were withdrawn because of study medication workflow errors and 1 was withdrawn by the surgeon.

**Fig 1.**
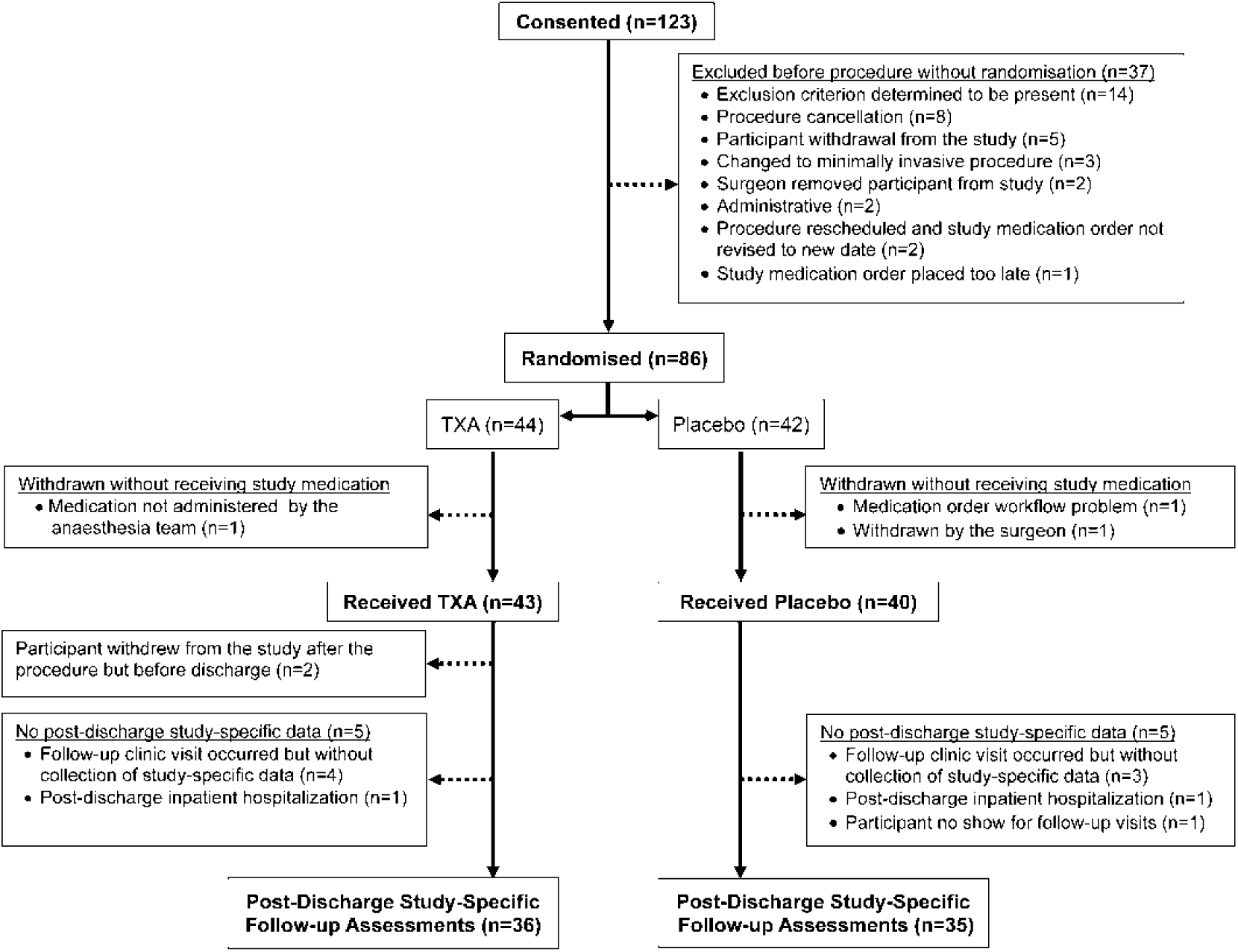
Flow diagram of participant enrollment, exclusion, randomization, dropout, and assessment.

Among 83 participants who received study medication (TXA [n=43]; Placebo [n 40]) all had consented to measurements of blood loss and transfusion. Because modifications to study outcomes (see Methods, Protocol and outcomes modifications) among these 83, 68 had also consented to postoperative delirium assessments and to preoperative and post-discharge cognitive testing, and 43 had also consented to pre- and post-operative blood collection. Two participants, both of whom received TXA, withdrew from the study prior to discharge; one on postoperative day 1 and the other on postoperative day 3. Five participants in each group who were eligible for post-discharge follow-up assessments did not receive study-specific assessments.

Based on *P* values <0.05, there was one statistically significant but clinically unimportant difference between groups in preoperative creatinine concentration (Table 1). In contrast, based on standardized mean differences >0.20, the TXA and Placebo groups differed in several characteristics. For example, although the median number of vertebral levels instrumented did not differ between groups (median=3 levels), the percentage of participants who underwent procedures with instrumentation of ≥5 levels was numerically greater in participants randomised to TXA (9/43=21%) *vs* Placebo (3/40=7.5%). The 43 participants in the TXA group had a total of 161 levels instrumentation *vs* 126 levels in the 40 Placebo participants.

**Table 1.** Participant, intraoperative, and postoperative characteristics. Data are expressed as median (25^th^, 75^th^ percentiles) or n (%). *Incision to wound closure.

| Variables | Tranexamic<br>Acid (n = 43) | Placebo<br>(n = 40) | P value | Standardized<br>mean difference |
| --- | --- | --- | --- | --- |
| Participant |  |  |  |  |
| Age, years | 59 (52, 72) | 63 (55, 71) | 0.494 | 0.153 |
| Female sex, n (%) | 20 (47%) | 22 (55%) | 0.512 | 0.168 |
| Weight, kg | 98 (78, 111) | 94 (80, 107) | 0.588 | 0.109 |
| Body mass index, kg m<sup>-2</sup> | 32 (28, 37) | 32 (28, 37) | 0.866 | 0.021 |
| Smoking, n (%) |  |  | 0.550 | 0.021 |
| Never | 16 (37%) | 17 (43%) |  |  |
| Former | 19 (44%) | 19 (48%) |  |  |
| Current | 8 (19%) | 4 (10%) |  |  |
| Hypertension, n (%) | 33 (77%) | 24 (60%) | 0.155 | 0.362 |
| Diabetes mellitus, n (%) | 9 (21%) | 11 (28%) | 0.609 | 0.152 |
| Preoperative creatinine, μmol L<sup>-1</sup> | 88 (71, 97) | 80 (63, 88) | **0.046** | 0.446 |
| Preoperative haemoglobin, g L<sup>-1</sup> | 140 (132, 148) | 133 (124, 145) | 0.059 | 0.434 |
| American Society of Anesthesiologists Class, n (%) |  |  | 0.386 | 0.298 |
| 1 | 1 (2%) | 1 (3%) |  |  |
| 2 | 31 (72%) | 22 (55%) |  |  |
| 3 | 10 (23%) | 16 (40%) |  |  |
| 4 | 1 (2%) | 1 (3%) |  |  |
| Intraoperative |  |  |  |  |
| Surgeon, n (%) |  |  | 0.722 | 0.450 |
| A | 5 (12%) | 5 (13%) |  |  |
| B | 3 (7%) | 6 (15%) |  |  |
| C | 4 (9%) | 4 (10%) |  |  |
| D | 1 (2%) | 0 (0%) |  |  |
| E | 10 (23%) | 5 (13%) |  |  |
| F | 12 (28%) | 10 (25%) |  |  |
| G | 8 (19%) | 10 (25%) |  |  |
| Procedure time*, minutes | 258 (226, 361) | 243 (195, 307) | 0.174 | 0.362 |
| Vertebral levels instrumented, n | 3 (2, 4) | 3 (2, 3) | 0.104 | 0.287 |
| Surgery time per vertebral level instrumented, minutes | 86 (74, 109) | 90 (76, 102) | 0.809 | 0.039 |
| Postoperative |  |  |  |  |
| Intensive care unit admission, n (%) | 5 (12%) | 3 (8%) | 0.714 | 0.141 |
| Length of hospital stay, days | 3 (2, 6) | 3 (2, 5) | 0.968 | 0.201 |
| Post-discharge disposition, n (%) |  |  | 0.136 | 0.457 |
| Home | 31 (72%) | 30 (75%) |  |  |
| Inpatient rehabilitation | 12 (28%) | 7 (18%) |  |  |
| Skilled care | 0 (0%) | 3 (8%) |  |  |

Bold character highlights *P* value <0.050.

### Blood loss and red blood cell transfusion

#### Blood loss

In linear robust regression models, intraoperative, postoperative, and total blood loss were each associated with the number of instrumented levels; all *P* values <0.002. For this reason, all blood loss results are reported with adjustment for the number of instrumented levels (Table 2).

**Table 2.** Blood loss and transfusion. Data are expressed as median (25^th^, 75^th^ percentiles) or n (%). * Four participants (TXA [n = 1]; Placebo [n = 3]) did not have postoperative wound drains and, for this reason, did not have postoperative blood loss measurements and did not have total (intraoperative plus postoperative) blood loss measurements.

| Variables | Tranexamic Acid (n = 43) | Placebo (n = 40) | *P* value |
| --- | --- | --- | --- |
| Blood loss per instrumented vertebral level, mL level<sup>-1</sup> |  |  |  |
| Intraoperative | 167 (87, 250) | 110 (67, 176) | 0.311 |
| Postoperative* | 128 (97, 186) | 207 (132, 258) | **0.013** |
| Total* | 305 (186, 412) | 333 (241, 428) | 0.472 |
| Patients receiving red blood cell transfusion |  |  |  |
| Intraoperative, n (%) | 7 (16%) | 6 (15%) | 0.873 |
| Postoperative, n (%) | 9 (21%) | 8 (20%) | 0.916 |
| Total, n (%) | 10 (23%) | 10 (25%) | 0.853 |

Bold character highlights *P* value <0.050.

Intraoperative estimated blood loss in the TXA and Placebo groups did not differ, with medians of 167 and 110 mL per level, respectively; *P*=0.311. Postoperative day 0 wound drain output was ∼37% less in participants who received TXA *vs* Placebo; (median [interquartile range]) 61 [48, 85] *vs* 97 [68, 140] mL per level, respectively, *P*=0.001. The duration of postoperative wound drain placement did not differ between the TXA and Placebo groups; 1 (1, 2) (median [interquartile range]) *vs* 1 (1, 2) days, respectively; *P*=0.258. Postoperative blood loss was ∼38% less in participants who received TXA compared with those who received Placebo, with medians of 128 and 207 mL per level, respectively; *P*=0.013, with no difference in TXA effect size between females (−0.61 [95% CI −1.20 to 0.04) and males (−0.68 [95% CI −1.30 to −0.03) (Supplementary data Table S2).

#### Red blood cell transfusion

In a logistic regression model, administration of ≥1 unit of red blood cells (RBCs) was associated with the number of instrumented vertebral levels (*P*=0.020), without a significant levels*group interaction (*P*=0.107). Percentages of participants who received ≥1 unit of RBCs did not differ between groups intraoperatively (15-16%), postoperatively (20-21%), or in total (23-25%); all *P* values ≥0.853, (Table 2). Additional transfusion results are provided in Supplementary data.

### Delirium

#### Delirium incidence

Overall delirium incidence was 18/65=28% (Table 3) In a logistic regression model of delirium incidence, there was no association with the number of instrumented levels (*P*=0.387) or a significant levels*group interaction (*P*= 0.779). Delirium incidence was numerically but not significantly less in participants who underwent instrumentation at ≤4 levels (13/55=24%) *vs* those who had instrumentation at ≥5 levels (5/10=50%); *P* =0.124. In other logistic regression models, delirium incidence was not associated with either intraoperative blood loss (*P*=0.472) or total blood loss (*P*=0.950).

**Table 3.** Delirium incidence. Data are expressed as n (%).

| **Delirium incidence** | **Tranexamic Acid**<br>**(n = 32)** | **Placebo**<br>**(n = 33)** | ***P* value** | **Odds ratio**<br>**(95% CI)** | **Effect size (h)**<br>**(95% CI)** |
| --- | --- | --- | --- | --- | --- |
| Overall | 7/32 (22%) | 11/33 (33%) | 0.408 | 0.56 (0.19 to 1.69) | -0.258 (-0.744 to 0.229) |
| Delirium incidence—By number of vertebral levels instrumented |  |  |  |  |  |
| ≤4 Levels | 4/25 (16%) | 9/30 (30%) | 0.341 | 0.45 (0.09 to 1.94) | -0.336 (-0.867 to 0.195) |
| ≥5 Levels | 3/7 (43%) | 2/3 (67%) | 1 | 0.41 (0.01 to 11.8) | -0.483 (-1.836 to 0.869) |
| Delirium incidence—By age |  |  |  |  |  |
| Older (≥65 years) | 3/10 (30%) | 9/17 (53%) | 0.424 | 0.38 (0.07 to 1.99) | -0.470 (-1.251 to 0.311) |
| Younger (<65 years) | 4/22 (18%) | 2/16 (13%) | 1 | 1.56 (0.25 to 9.75) | 0.158 (-0.486 to 0.802) |
| Delirium incidence—By sex |  |  |  |  |  |
| Males | 3/17 (18%) | 5/14 (36%) | 0.413 | 0.39 (0.07 to 2.03) | -0.414 (-1.121 to 0.293) |
| Females | 4/15 (27%) | 6/19 (32%) | 1 | 0.79 (0.18 to 3.53) | -0.104 (-0.785 to 0.569) |

Delirium incidence was numerically but not significantly less in the TXA group (7/32 = 22%) *vs* Placebo (11/33=33%); *P*=0.408. Delirium incidence was greater in older (12/27=44%) *vs* younger participants (6/38=16%); *P*=0.023. Among older participants, delirium incidence was numerically but not significantly less in the TXA group (3/10=30%) *vs* Placebo (9/17=53%); *P*=0.424. Among younger participants, delirium incidence was not significantly greater in the TXA group (4/22=18%) *vs* Placebo (2/16=13%). Delirium incidence did not differ between males (8/31=26%) and females (10/34=29%); *P*=0.788. Among males, delirium incidence was numerically but not significantly less in the TXA group (3/17=18%) vs placebo (5/14=36%); *P*=0.413. Among females, delirium incidence was not significantly less in the TXA group (4/15=27%) vs Placebo (6/19=32%); *P*=1; see Discussion, Delirium Clinical Trial Implications.

Participants who had delirium (n=18) had a greater length of stay than those who did not (n=47); 7 (5, 8) *vs* 3 (2, 4) days, respectively; *P=*0.000004, and were less often discharged to home; (8/18=44%) *vs* (38/47=83%), respectively; *P*=0.006.

#### Delirium severity

In a robust linear regression model that included the number of instrumented levels and group (TXA *vs* Placebo) as the independent variables and maximum delirium severity as the dependent variable, there was a positive association between the number of instrumented levels and maximum delirium severity in the Placebo group (n=33: *P*=0.001), with a non-significant negative levels*group interaction term (Beta= −0.40 [95% CI −0.94 to 0.15]; *P*=0.161) (Fig 2a). In other robust linear models, maximum delirium severity was not associated with either intraoperative blood loss (*P*=0.316) or total blood loss (*P*=0.659), with no significant levels*group interaction terms (all *P* values ≥0.528)

**Fig 2.**
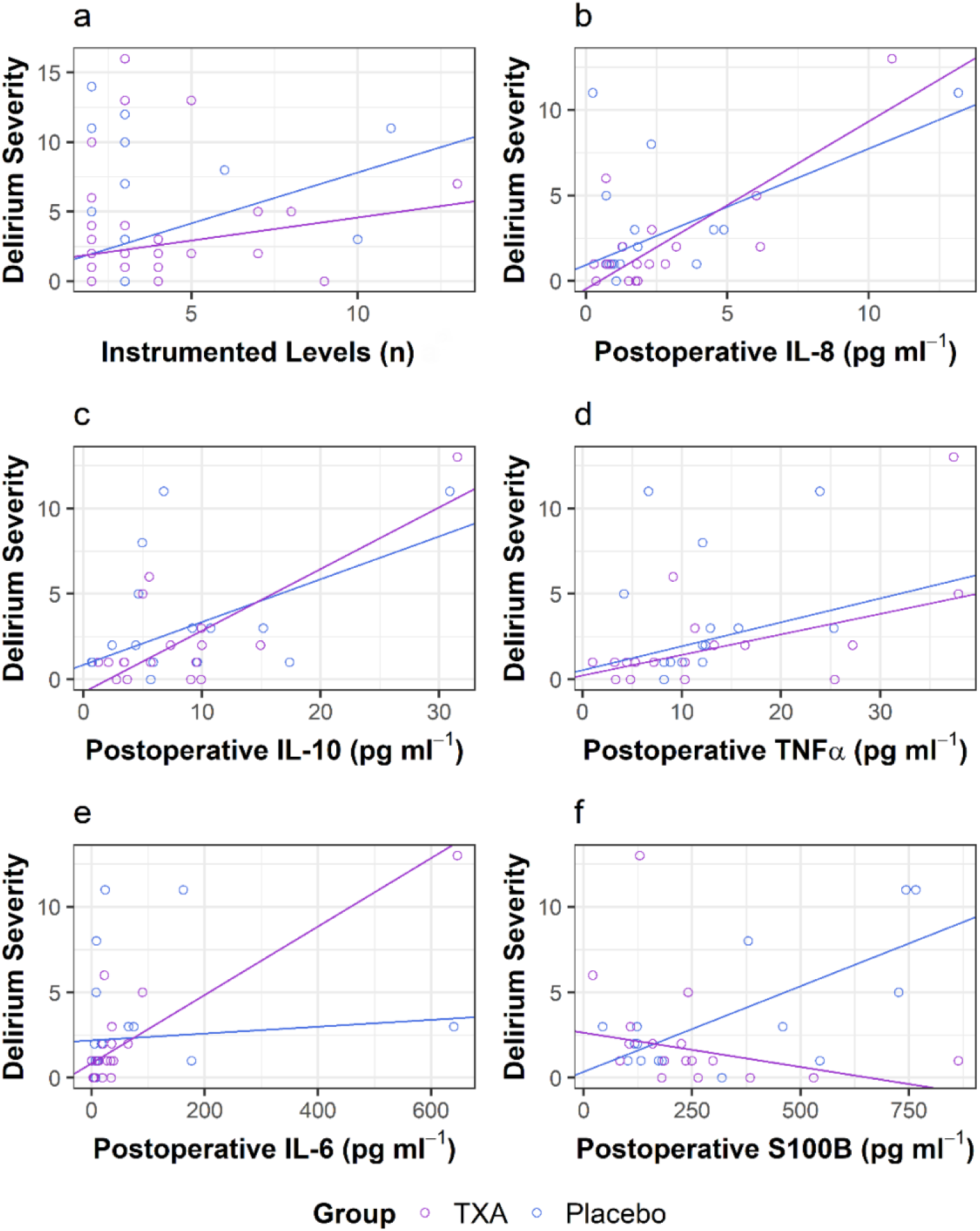
Maximum postoperative delirium severity *vs* a) number of instrumented vertebral levels and *vs* postoperative concentrations of b) IL-8; c) IL-10; d) TNFα; e) IL-6; and f) S100B. TXA data points are shown in purple circles and lines and Placebo data points are shown in blue circles and lines. In panel a) TXA (n=32) and Placebo (n=33). In panels b-f) TXA (n=16) and Placebo (n=16). Robust regression lines are shown.

In a robust linear regression model that included maximum postoperative delirium severity and postoperative delirium diagnosis (no [n=47] vs yes [n=18]) as the independent variables and length of stay as the dependent variable, there was a positive association in the no delirium group (*P*=0.0000002) with a non-significant severity*delirium group interaction term (*P*=0.471) (Fig3). This indicates the association between delirium severity and length of stay did not depend on whether participants met the diagnostic criteria for delirium; see Discussion, Delirium incidence and severity. Additional delirium severity results are provided in Supplementary data, Table S3.

### Biomarkers of systemic inflammation and neuroinflammation

#### Preoperative and postoperative biomarker concentrations

Preoperative concentrations of each of the five biomarkers did not differ between the TXA and Placebo groups (all *P* values ≥0.418). In both groups, postoperative concentrations of 3 of the 5 biomarkers were significantly greater than corresponding preoperative concentrations (IL-10, IL-6, and S100B; all *P* values ≤0.010) (Table 4). In the TXA group, TNFα decreased postoperatively (*P*=0.006). Postoperative concentrations of each of the five biomarkers did not differ between the two groups (all *P* values ≥0.711).

**Table 4.** Plasma biomarker concentrations. Data are expressed as median (25^th^, 75^th^ percentiles). Only values from samples with paired pre- and post-operative values (n=31) that were used to calculate the *P* values are shown.

| **Biomarkers** |  | **Tranexamic Acid**<br>**(n = 16)** | **Placebo**<br>**(n = 15)** | **Overall**<br>**(n = 31)** |
| --- | --- | --- | --- | --- |
| Interleukin-8, pg mL<sup>-1</sup> |  |  |  |  |
|  | Preoperative | 1.40 (0.95, 1.80) | 1.38 (0.96, 1.95) | 1.38 (0.93, 1.93) |
|  | Postoperative | 1.79 (0.80, 2.55) | 1.30 (0.95, 3.11) | 1.73 (0.87, 2.77) |
|  | *P* value | 0.105 | 0.590 | 0.134 |
| Interleukin-10, pg mL<sup>-1</sup> |  |  |  |  |
|  | Preoperative | 2.25 (1.30, 3.42) | 2.93 (1.58, 3.99) | 2.31 (1.46, 3.84) |
|  | Postoperative | 5.59 (3.43, 9.91) | 5.86 (4.50, 10.16) | 5.65 (3.56, 9.92) |
|  | *P* value | **0.003** | **0.010** | **0.00004** |
| Tumor Necrosis Factor, alpha, pg mL<sup>-1</sup> |  |  |  |  |
|  | Preoperative | 14.91 (8.59, 20.10) | 13.13 (9.77, 15.84) | 14.17 (9.29, 16.61) |
|  | Postoperative | 9.70 (4.72, 18.61) | 12.10 (8.21, 12.62) | 10.29 (5.97, 14.49) |
|  | *P* Value | **0.006** | 0.083 | **0.002** |
| Interleukin-6, pg mL<sup>-1</sup> |  |  |  |  |
|  | Preoperative | 1.32 (0.79, 3.58) | 1.34 (0.56, 2.00) | 1.34 (0.72, 2.80) |
|  | Postoperative | 28.22 (11.70, 36.81) | 10.96 (7.78, 70.01) | 20.70 (8.32, 51.47) |
|  | *P* value | **0.0002** | **0.0001** | **<0.0000001** |
| S100B, pg mL<sup>-1</sup> |  |  |  |  |
|  | Preoperative | 118.8 (69.6, 174.8) | 116.6 (73.3, 150.5) | 117.4 (68.3, 163.5) |
|  | Postoperative | 229.9 (151.0, 271.9) | 180.5 (123.1, 501.6) | 224.7 (126.1, 381.5) |
|  | *P* value | **0.005** | **0.0002** | **0.000003** |

Bold characters highlight *P* values <0.050.

#### Postoperative biomarker concentrations per vertebral levels instrumented

In robust linear regression models that included the number of instrumented levels and group (TXA *vs* Placebo) as the independent variables and postoperative cytokine concentrations as the dependent variable, in the Placebo group (n=16) postoperative cytokine concentrations (IL-8, IL-10, TNFα, and IL-6) were each positively associated with the number of instrumented vertebral levels; all *P* values <0.007. In contrast, S100B was not associated with the number of instrumented levels, either in the Placebo group (n=16: *P*=0.284) or overall (n=32: *P*=0.093). In all four cytokine models and in the S100B models, there were no significant level*group interaction terms (all *P* values >0.093), indicating that TXA did not significantly affect the associations between the number of instrumented levels and postoperative biomarker concentrations.

#### Cytokine/S100B interactions

With the possible exception of IL-8, there were no significant associations between postoperative cytokine concentrations and S100B concentrations and no effect of TXA on those associations; see Supplementary data, Fig S1.

#### Postoperative biomarkers and delirium incidence and severity

Postoperative TNFα concentrations were marginally but not significantly greater in participants who had postoperative delirium (n=7: 15.71 [10.60, 30.66] pg mL^-1^) *vs* those who did not (n=25: 10.03 [5.30, 12.36] pg mL^-1^); *P*=0.055. Postoperative IL-6 concentrations were significantly greater in participants who had postoperative delirium (n=7: 90.03 [23.35, 401.01] pg mL^-1^) *vs* those who did not (n=25: 17.67 [7.31, 35.47)] pg mL^-1^); *P*=0.026.

In robust linear regression models, in the Placebo group (n=16), delirium severity was positively associated with postoperative IL-8 (*P*=0.00008; Fig 2b) and IL-10 (*P*=0.005; Fig 2c) and was positively but not significantly associated with postoperative TNFα (*P*=0.079; Fig 2d), with no significant cytokine*group interaction terms (all *P* > 0.203) indicating that TXA did not affect these associations. In contrast, with IL-6, in the Placebo group there was no association with delirium severity (*P*=0.520) but there was a significant positive IL6*group interaction term (Beta=0.018 [95% CI 0.01 to 0.025] *P*=0.00004^1^; Fig 2e). This indicates that delirium severity associated with IL-6 was significant only in the TXA group; see Discussion. Finally, in the Placebo group, there was an association between S100B and delirium severity (*P* = 0.0009) with a significant negative S100B*group interaction term (Beta = −0.01 [95% CI −0.02 to −0.01]; *P*=0.002; Fig 2f) indicating that, compared with Placebo, TXA significantly decreased delirium severity associated with S100B; see Discussion, Mechanistic hypotheses regarding delirium and TXA.

#### Cognitive Testing

Preoperative scores of each of the 4 cognitive tests did not differ between groups (all *P* values ≥0.598). For both the TICSm and TMT-A tests, post-discharge test scores did not significantly differ from preoperative scores in either group or overall (all *P* values ≥0.065), (Table 5). In contrast, there was an overall post-discharge improvement in TMT-B test scores (*P*=0.013); the *more negative* post-discharge *z*-scores indicating participants required *less time* to complete the test. Improvements in post-discharge TMT-B scores were significant in participants who received TXA (*P* = 0.007) but were not significant in the Placebo group (*P*=0.303) Likewise, there was an overall post-discharge improvement in the COWA *z*-score (*P*=0.0007); *less negative* postoperative COWA *z*-scores indicating participants provided a *greater number* of words. COWA scores significantly improved in the TXA group (*P*=0.009), whereas COWA scores significantly *worsened* in the Placebo group (*P*=0.033).

**Table 5.**
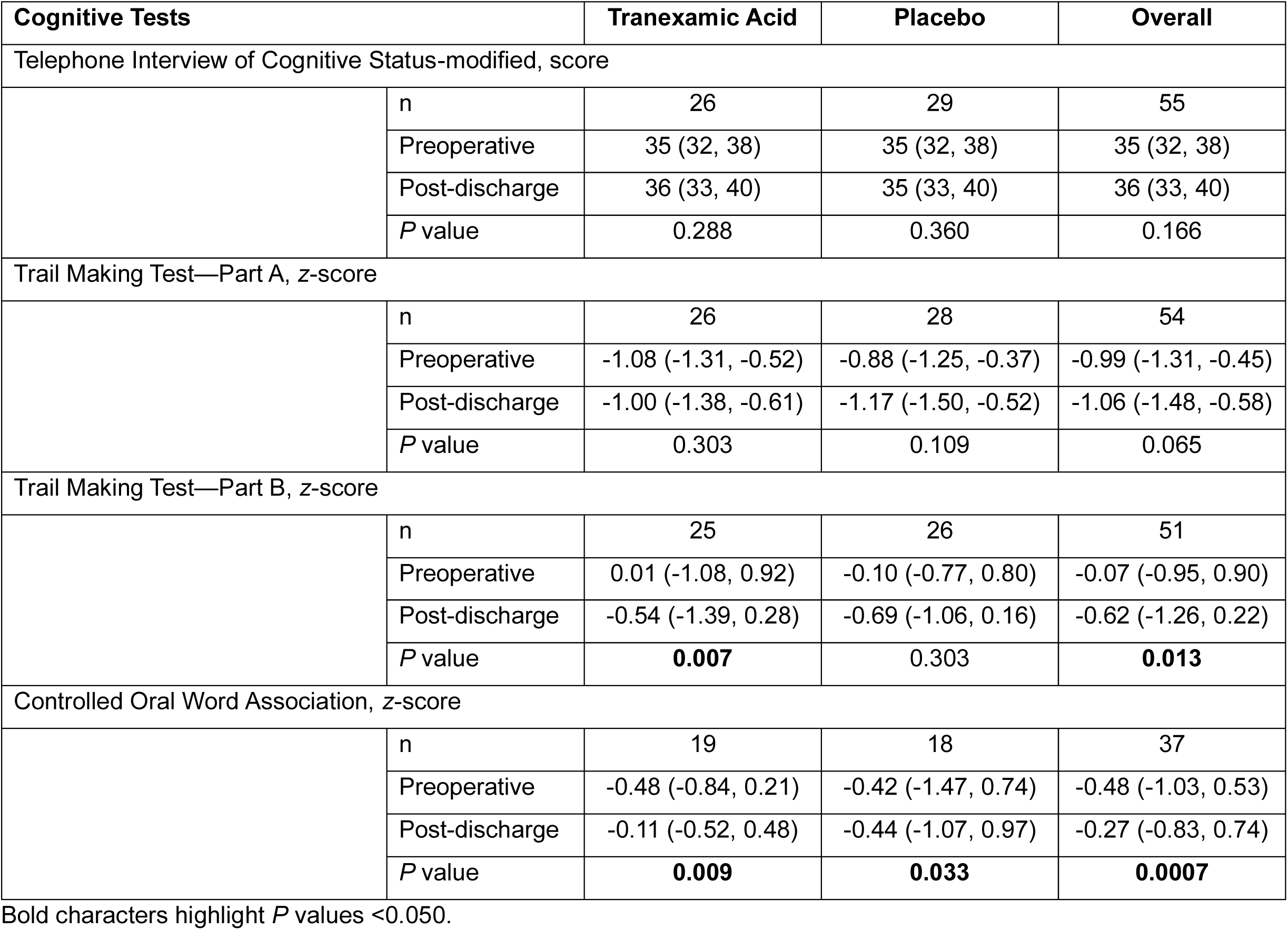
Cognitive test scores. Data are expressed as median (25^th^, 75^th^ percentiles).

In participants who experienced postoperative delirium, there were no differences between preoperative and post-discharge test scores (all *P* values ≥0.210). In contrast, in participants without delirium, there was a marginal but non-significant post-discharge improvement in TMT-A scores (*P*=0.055) and signficiant improvements in post-discharge TMT-B (*P* =0.0004) and COWA (*P* =0.0006) scores (Supplementary data.Table S4).

#### Adverse Events

The two groups did not differ in the incidence of adverse events (Table 6). Although 8 participants had vision-related complaints within 3 months after discharge (TXA [n = 7], Placebo [n = 1]), 3 were due to pre-existing conditions, 2 were due to orthostatic hypotension, and 1 was due to a corneal abrasion. The 2 remaining participants, both of whom had received TXA, had vision complaints on postoperative days 7 and 82 that were unexplained and were temporary.

**Table 6.** Adverse events through final post-discharge follow up. Data are expressed n (%). * Pulmonary embolism 22 days after procedure (n=1); embolic stroke 69 days after procedure (n=1). ‡Unilateral vision change on postoperative day 7 (n=1); worsening vision occurred briefly on postoperative day 82 (n=1) with no subsequent reports regarding vision with either participant.

| **Adverse Event** | **Tranexamic Acid**<br>**(n=43)** | **Placebo**<br>**(n=40)** | **P Value** |
| --- | --- | --- | --- |
| Thromboembolic event | 0 (0%) | 2 (5%)* | 0.229 |
| Vision abnormality | 2 (5%)‡ | 0 (0%) | 0.495 |
| Renal dysfunction on postoperative day 1 | 2 (5%) | 4 (10%) | 0.422 |
| Wound complication | 2 (5%) | 2 (5%) | 1 |
| Seizure | 0 (0%) | 0 (0%) | 1 |
| Death | 0 (0%) | 0 (0%) | 1 |

## Discussion

This study has a small sample size; hence, it is possible that clinically or mechanistically important effects of TXA were not detected. In addition, because of multiple comparisons it is likely that some of the reported effects of TXA are false positives. Accordingly, we consider this study’s findings to be valuable primarily for hypothesis generation. We propose that the potential effect TXA to decrease delirium incidence by 33% is clinically important and is sufficient to justify an adequately powered clinical trial.

### Blood loss and TXA concentrations

In this study, although TXA did not decrease *intra*operative blood loss, TXA significantly decreased *post*operative blood loss by ∼38%. These findings are compatible with those of a recent observational report of patients who underwent 1-3 level transforaminal lumbar interbody fusion and who received TXA (10 mg kg^-1^) prior to incision.^47^ TXA did not decrease intraoperative blood loss but decreased postoperative wound drain output by 20-30%.^47^

In the current study, the apparent lack of effect of TXA on intraoperative blood loss may be explained by multiple potential sources of error in intraoperative blood loss estimates.^48,49^ In contrast, all postoperative blood loss was collected in a single container (wound drains), was not diluted with any fluid, and volumes were objectively measured rather than estimated. Thus, postoperative blood loss volumes are more likely to be accurate. It is also plausible the effect of TXA to decrease blood loss may be proportionately less *intra*operatively than *post*operatively.

TXA decreases blood loss by inhibition of fibrinolysis, promoting clot stability. Intraoperatively, spine surgeons do not rely solely on clot stability to control intraoperative bleeding. Instead, surgeons directly observe the sources of bleeding and apply one or more active interventions to decrease it (*e.g*., electrocautery). The more effective these active measures are, the less effect TXA should have on intraoperative blood loss. In contrast, *post*operatively, there are no active measures taken to decrease bleeding and blood loss will be determined only by the balance of clot formation and clot lysis. Accordingly, TXA may have greater potential to decrease *post*operative bleeding—both absolutely and relatively—than *intra*operative bleeding.

The 38% decrease in postoperative blood loss in this study is consistent with the effect of TXA reported in multiple randomised control trials of patients undergoing posterior lumbar interbody fusions, with an average of a 33% reduction (range 20% to 47%).^18^ It is also consistent with a metaanalysis of TXA studies in adults undergoing cardiac procedures in which the maximum effect of TXA on blood loss reduction was 40% (95% credible interval 34% to 47%).^16^

*In vitro* studies indicate TXA concentrations of 10-15 mg L^-1^ result in 80% inhibition of plasmin-mediated fibrinolysis.^50^ In the aforementioned meta-analysis of adult cardiac procedures, the systemic TXA concentration necessary to achieve 80% of the maximum decrease perioperative blood loss was estimated to be 22.4 mg L^-1^ (95% CI ∼10 to 40 mg L^-1^).^16^ In a pharmacokinetic study of older patients undergoing elective hip arthroplasty, Lanoiselée and colleagues used a TXA dosing regimen (∼13 mg kg^-1^ loading dose; ∼1.8 mg kg^-1^ h^-1^ infusion) that was nearly identical to that used in this study (10 mg kg^-1^ loading dose; 2 mg kg^-1^ h^-1^ infusion).^51^ The Lanoiselée dosing regimen resulted in a steady-state TXA plasma concentration of ∼30 mg L^-1^, which is substantially greater than the TXA concentration needed for near maximum inhibition of plasmin-mediated fibrinolysis. Using pharmacokinetic modeling, Lanoiselée and colleagues estimated TXA concentrations would exceed 10 mg L^-1^ for approximately 3 hours after cessation of the infusion in patients who had normal renal function. Therefore, the intraoperative TXA dosing regimen used in our study would have resulted in clinically effective TXA concentrations (∼30 mg L^-1^) intraoperatively and for several hours thereafter. The fact that, in our study, TXA decreased blood loss measured *after* discontinuation of the infusion provides strong indirect evidence that our TXA dose was sufficient to inhibit plasmin. As will be discussed, *brain* plasmin inhibition is hypothesized to be the mechanism by which TXA may decrease delirium.

### Delirium incidence and severity

Delirium incidence in the TXA group (7/32=22%) was numerically but not significantly less than in the Placebo group (11/33=33%); *P*=0.408, absolute difference = 11%, relative difference = 33%, effect size = −0.25<u>8</u> (95% CI −0.744 to 0.229). This potential effect of TXA to decrease delirium incidence is consistent with a 33% relative decrease in delirium incidence reported in our prior retrospective observational study using a propensity score-based analysis; TXA (14%) *vs* controls (21%); *P*=0.004.^45^ In addition, in this study, participants who received TXA had significantly improved post-discharge performance in 2 of 4 cognitive tests (TMT-B [*P* = 0.007]; COWA [*P* = 0.009]) compared to their preoperative baselines, whereas those who received Placebo did not improve in three of the four tests and worsened in one. This suggests that, in addition to potentially decreasing postoperative delirium, TXA has the potential to favorably affect longer-term cognitive outcomes. Furthermore, participants who did not have postoperative delirium had significantly improved performance in 2 of 4 post-discharge cognitive tests (TMT-B [*P*=0.0004], COWA [*P*=0.0006]), whereas those who had delirium did not improve in any of the tests. This observation is consistent with the linkage between postoperative delirium and long-term cognitive outcomes.^5,6^ A unifying hypothesis would be that, by decreasing the incidence of postoperative delirium, TXA may also decrease long-term postoperative cognitive dysfunction.

We observed a significant association between maximum postoperative delirium severity and length of stay that was independent of whether patients met the formal diagnostic criteria for delirium. This suggests the behavioural and cognitive abnormalities included in the 3D-CAM, even when not sufficient to meet the diagnostic criteria for delirium (*i.e*., “subclinical” delirium), adversely affect postoperative recovery. In this study, decreasing maximum delirium severity by ∼2 points (out of 20) would be predicted to decrease length of stay by 1 day (Fig 3). Thus, both delirium incidence and delirium severity are clinically important.

**Fig 3.**
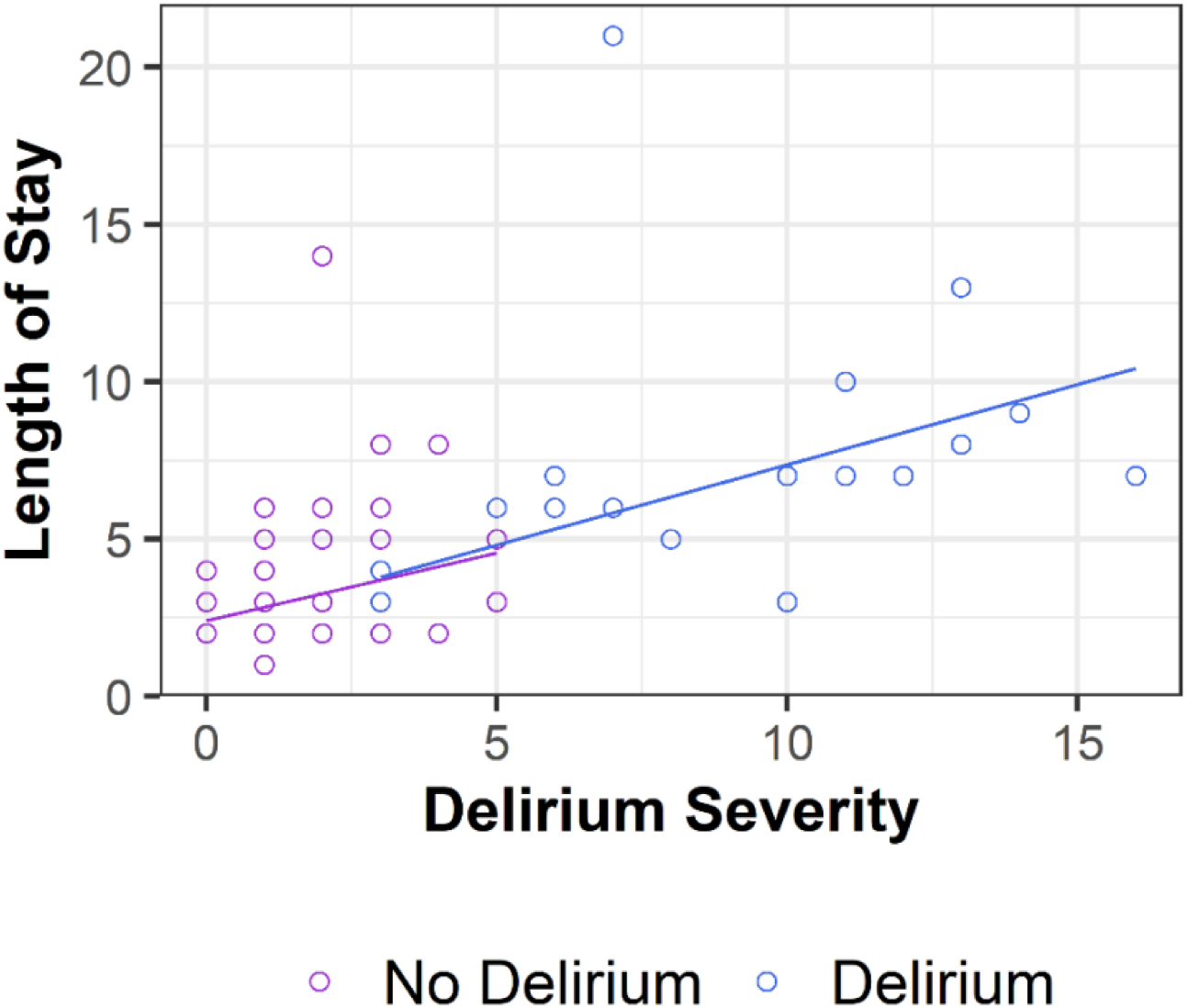
Postoperative length of stay *vs* maximum postoperative delirium severity. Participants who did not meet the diagnostic criteria for delirium (n=47) are shown in purple circles and lines. Participants who met the diagnostic criteria for delirium (n=18) are shown in blue circles and lines. Robust regression lines are shown.

### Mechanistic hypotheses regarding delirium and TXA

#### Cytokines

In murine models of abdominal^52^ and orthopedic procedures,^13^ and burns,^53^ systemic IL-6^13,52,53^ and IL-1β^52^ mediate increased blood-brain barrier permeability, which occurs within 3-6 h after injury. Other procedure-induced systemic proinflammatory mediators (*e.g*., TNFα^54^, IL-17^55^, high mobility group box protein 1 (HMGB1)^56^, and others^57^) also contribute to postoperative blood-brain-barrier dysfunction and neuroinflammatory delirium.

In a prospective observational study of patients who underwent elective total hip or knee replacement, blood brain permeability (CSF/serum albumin ratio) and CSF concentrations of IL-6, IL-8, and S100B were each increased above preoperative baseline within 4 hours after incision.^58^ Two recent clinical studies reported significant associations between postoperative delirium and increased blood-brain barrier permeability to albumin 24 hours postoperatively.^28.59^ Because albumin has a molecular weight of 67,000 g mol^-1^ (67 kDa), it seems likely that both early and later (24 h) postoperative increases in blood-brain barrier permeability would permit CSF entry of systemic cytokines (*e.g.*, IL-6 [21 kDa], TNFα [26 kDa]) and/or other proinflammatory proteins (*e.g.*, HMGB1 [25-30 kDa], plasmin [90 kDa]), thereby either initiating or increasing neuroinflammation^28,60^ with consequent electrophysical^61,62^ and cognitive abnormalities.

In our study, in participants randomised to Placebo we observed: 1) a positive association between the number of instrumented levels and delirium severity (Fig 2a); 2) positive associations between the number of instrumented levels and 24 hour cytokine concentrations; and 3) positive associations between 24 hour cytokine concentrations and delirium severity (Fig 2b-e). These observations are consistent with the animal and clinical literature that indicate postoperative delirium occurs, at least in part, as the result of systemic inflammatory responses determined by the magnitude of tissue injury. With a few exceptions and qualifiers, these associations were not affected by intraoperative TXA.

Although intraoperative TXA did not significantly decrease the positive association between the number of instrumented levels and postoperative delirium severity (levels* group interaction Beta = −0.40 [95% CI −0.94 to 0.15]; *P*=0.161; Fig 2a), statistical significance might be achieved with a larger sample size. Although intraoperative TXA administration did not significantly affect cytokine concentrations 24 hours postoperatively, two factors likely explain this observation. First, because the elimination half-lives of these four cytokines are brief (IL-8: ∼5 min^63^; IL-10: ∼3 h^64^; TNFα: 15-30 min^65^; IL-6: <30 min^66,67^), systemic cytokine concentrations at 24 hours postoperatively likely reflect ongoing synthesis at the surgical site with subsequent entry into the systemic circulation. Second, because TXA has an elimination half-life of 2-3 hours in patients who have normal renal function,^68^ TXA administered only intraoperatively should be entirely eliminated by 24 h postoperatively. In combination, these two factors predict that TXA administered only intraoperatively (this study) should not affect systemic cytokine concentrations 24 hours postoperatively.

In this study, compared with Placebo, intraoperative TXA administration did not significantly decrease the strength of the association between postoperative IL-8, IL-10, or TNFα concentrations and delirium severity. The difference between the Placebo and TXA groups in their associations between IL-6 and delirium severity could be easily misinterpreted as indicating that TXA increased delirium severity. However, the difference between groups is present because the Placebo group exhibited no association between IL-6 and delirium severity—which differs from the behavior of the three other cytokines in this study and is quite inconsistent with the literature. Most likely, the attenuated delirium response to IL-6 in the Placebo group is an artifact of small sample size. To the extent that TXA may potentially decrease delirium incidence or severity, our data suggest TXA does so by means other than affecting systemic cytokines and/or their effects on the brain.

#### S100B

S100B is a homo-dimeric protein (molecular weight 21 kDa) that is secreted primarily by astrocytes in response to a wide variety of pro-inflammatory stimuli.^69^ Once secreted into brain extracellular fluid, S100B has numerous intra- and extra-cellular targets (*e.g*., the receptor for advanced glycated end products [RAGE]^69^), some of which result in increased blood-brain barrier permeability. S100B can enter the systemic circulation via the altered blood-brain barrier and/or via the glymphatic system.^69,70^ Because in the systemic circulation S100B elimination half-life is ∼1 hour,^71^ systemic S100B concentrations 24 hours postoperatively likely reflect ongoing astrocytic secretion and subsequent entry into the systemic circulation.

Several prior observational studies^28,72–74^ have reported associations between S100B and either delirium incidence or severity, which suggests that astrocyte activation (and/or blood-brain barrier dysfunction) is involved in (or is coincident with) the pathophysiology of delirium.

Consistent with these prior studies, in the Placebo group there was a strong positive association between S100B and delirium severity (Fig 2e). In contrast, in the TXA group, the association between S100B and delirium severity appeared to be entirely eliminated (Fig 2e). This latter observation supports the hypothesis that TXA’s potential effect to decrease delirium takes place downstream of astrocyte activation and S100B secretion.

#### The potential role of brain plasmin or plasminogen activators

A biologically plausible mediator of postoperative delirium—which is inhibited by TXA—is *brain* plasmin(ogen). In animal and *in vitro* models, plasmin mediates neuroinflammation *via* multiple mechanisms,^22,75^ contributing to increased blood-brain barrier permeability, degradation of extracellular matrix proteins, leukocyte diapedesis, and activation of brain immune cells (astrocytes and microglia) to secrete inflammatory cytokines.^76^

In humans, the transfer of TXA from blood to CSF normally requires many hours.^77–80^ However, because operative procedures result in a very early postoperative increase in blood-brain permeability,^58^ it seems likely blood-brain transfer of small molecules like TXA (0.157 kDa) would occur more rapidly than normal. If TXA gains rapid access to the brain in this way, by inhibiting plasmin(ogen) binding to its receptors, TXA may decrease postoperative neuroinflammation and consequent delirium.^19,20,23,75^

In three reports using murine models of traumatic brain injury, a single post-insult dose of intravenous TXA normalized blood-brain-barrier permeability,^26,81^ brain cytokine concentrations,^82^ and improved functional^26,81^ and histologic^82^ outcomes. Notably, in one of these three studies, although TXA *increased* some *systemic* cytokine concentrations, it *decreased* brain cytokine concentrations ^82^ Thus, TXA’s beneficial effect did *not* require a decrease in systemic cytokine concentrations. In another of these three studies, that of Daglas and colleagues, although TXA’s beneficial effects on blood-brain barrier permeability and functional outcome were shown to be plasmin dependent, these benefits occurred only in male mice.^26^ The authors reported: 1) there were sex-dependent differences in brain plasminogen activation (tissue plasminogen activator [t-PA] in males; urokinase plasminogen activator [u-PA] in females); and 2) TXA had different effects on these two activators—although TXA decreased t-PA activity (decreasing plasmin), TXA *increased* u-PA activity (increasing plasmin). This finding suggests the potential benefit of TXA on postoperative delirium could be sex-dependent, with a greater benefit in males.

#### Antagonism of γ-aminobutyric acid receptors

Finally, another potential mechanism by which TXA may decrease delirium is by its stimulant properties, which may counteract the electrophysiologic inhibition that is hypothesized to occur in delirium.^83^ A recent study reported increased inhibitory interneuron activity in modelling of auditory evoked responses in patients with delirium.^84^ Acting through antagonism of γ-aminobutyric acid receptors,^80,85^ TXA may directly counter these inhibitory effects, decreasing the severity of delirium. However, given the rapid clearance of TXA from the circulation and CSF,^80^ this seems unlikely to be a mechanism by which TXA could decrease delirium beyond the first few hours after the procedure.

### Delirium clinical trial implications

#### Sex

Consistent with the aforementioned preclinical study of Daglas and colleagues,^26^ in this study TXA appeared to decrease delirium incidence to a greater extent in males (effect size −0.414) than in females (effect size −0.104). In contrast, the effect of TXA to decrease postoperative bleeding did not differ between the sexes (effect size ∼ −0.64; Supplementary data Table S2) This suggests TXA’s effect to inhibit plasmin(ogen) may differ between the brain and the periphery. We suggest that in a future clinical trial to test TXA’s effect on postoperative delirium randomization should be stratified by sex.

#### Age

Increasing patient age is well established as a risk factor for postoperative delirium.^7^ The relatively high incidence of postoperative delirium in older participants observed in this study (44.4%) is consistent with other studies in older spine fusion patients; 40.5%^86^ and 43.2%.^87^ A recent study reported numerous age-dependent differences in the postoperative delirium proteome.^88^ Although some proteins linked to delirium were common to both younger (45-60 years) and older (≥70 years) patients (*e.g*., IL-8 and IL-6), older patients had a greater number of delirium-associated proteins and pathways than younger patients. In addition to these systemic differences, neuroimmune responses change with age.^89^ For example, in murine models of abdominal procedures^52^ and traumatic brain injury,^90^ older animals had a more robust neuroinflammatory response than younger animals. This finding suggests the potential benefit of TXA on postoperative delirium could be age-dependent, with a greater benefit in older patients. Data from this study are consistent with that possibility; TXA appeared to decrease delirium incidence to a greater extent in older participants (effect size= −0.470) than in younger participants (effect size=0.158). Accordingly, we suggest that in a future clinical trial to test TXA’s effect on postoperative delirium, enrollment should be limited to older patients or randomization be stratified by age.

#### Adverse Events

In this study, the incidence of adverse events did not differ between groups but, because of its small sample size, the statistical power to detect TXA-related adverse events was very low. However, there are numerous large clinical trials and meta-analyses reporting the adverse event profile of perioperative TXA administration.

In a meta-analysis of 8 randomised controlled trials of TXA in patients undergoing posterior lumbar interbody fusion (the procedure in this study), there was no difference in the incidence of deep vein thrombosis between patients receiving TXA vs controls (10.5% *vs* 12.2%, respectively; *P*=0.33, OR=0.83 [95% CI 0.56 to 1.21]).^18^ In a recent randomised trial of TXA in 9,535 patients undergoing non-cardiac procedures, there were no differences between groups in the incidences of postoperative deep vein thrombosis, pulmonary embolism, seizure, acute kidney injury, infection, myocardial infarction, stroke, or mortality^17^ In a meta-analysis of 192 randomised controlled trials of intravenous TXA in 68,118 surgical patients, TXA did not increase the risk of venous thrombosis, pulmonary embolism, myocardial ischemia or infarction, or cerebral ischemia or infarction.^91^

As of March 31, 2024, the US Food and Drug Administration (FDA) Adverse Events Reporting System reports eye disorders constituted 130 of 3138 (4.1%) TXA adverse event reports since 1987.^92^ Among eye disorders, the four most common were visual impairment (n=29, 0.92%), retinal artery occlusion (n=18, 0.57%), blindness (n=17, 0.54%) and vision blurred (n = 16, 0.51%). The authors are aware of 2 prior reports of possible ophthalmologic adverse events associated with perioperative TXA administration; 1 case of temporary color distortion (dyschromatopsia)^93^ and 2 cases of postoperative central retinal arterial occlusion in 662 patients undergoing prone craniofacial procedures.^94^ The authors of the latter study did not ascribe the retinal arterial occlusions to TXA. In this study, all potential participants were screened and excluded for color vision defects and for any history of retinal vein or artery occlusion. Although 8 of 83 (10%) participants had at least 1 vision-related complaint within 3 months of follow-up (TXA [n=7], Placebo [n=1]), only 2 (both of whom received TXA) had unexplained but temporary visual symptoms. Although we cannot rule out the possibily that TXA contributed to these events, their onset on postoperative days 7 and 82 make this seem less likely.

TXA can cause seizures but, practically always, this requires the combination of both low TXA clearance (*i.e.*, low creatinine clearance) and large (>50 mg/kg intravenous bolus) and/or sustained TXA dosing.^95^ The authors are aware of 2 case reports of TXA-associated seizures in adults after non-cardiac procedures, one after a meningioma resection (TXA dose=44 mg kg^-1^; creatine clearance ≅35 mL min^-1^)^96^ and one after a 3-level lumbar fusion (TXA dose=61 mg kg^-1^; renal function not reported).^97^

### Limitations

This trial has numerous operational limitations, the first among them being that, among 123 consenting participants, 37 (30%) were excluded prior to their procedures without being randomised. This high rate of pre-randomization exclusion is similar to that reported in another recent study of postoperative delirium in adult spine procedures (25/124=20%).^98^ In our study, the interval between enrollment and the procedure was usually 2-4 weeks, during which time additional data from outside healthcare providers was received, new preoperative studies were conducted, and continued procedural planning occurred. The new information and/or plan often changed participants’ eligibility.

Another operational limitation was related to our center’s electronic medical record and the required workflow for physicians to enter orders for study medication. The system did not readily adjust for changes in the scheduled day of the procedure, As a consequence, some participants did not have study medications prepared when they otherwise should have— decreasing the number of consenting participants who were randomised.

Finally, although the basic intervention of this trial remained unchanged, there were numerous changes in outcome measures and their related procedures throughout the course of this trial. Additional challenges and opportunities for errors in study implementation were introduced because of a repeated loss of experienced study personnel. Lack of continuity and familiarity with protocol details contributed to incomplete data collection for many outcome measures. Missing data points decrease the information provided by the participants’ service and, by decreasing statistical power, decrease both the ability to detect adverse events and the reliability of findings. Although we stopped the trial early because our planned enrollment size was underpowered to detect the potential benefit of TXA on delirium, all of these other factors influenced our decision to stop enrollment. We are now using the knowledge gained and lessons learned from conducting this study to design a new protocol and refine the organizational structure of the research team in order to maximize the probability of definitively answering the scientific questions that this study has posed.

## Conclusion

Although not statistically significant, delirium incidence in the TXA group (22%) was numerically less than in the Placebo group (33%); absolute difference=11%, relative difference=33%, effect size = −0.25<u>8</u> (95% CI −0.744 to 0.229). This potential effect of TXA to decrease delirium is consistent with a 33% relative decrease in delirium incidence reported in a prior retrospective observational study in a similar population.^45^ The safety of intraoperative TXA at the dose used in this study has been established in numerous prior studies. Because of the consistency and size of the potential treatment effect of TXA to decrease postoperative delirium, a definitive randomised clinical trial is justified.

## Authors’ contributions

Conceptualization: MZ, RDS, MAH

Data curation: BJH, RWW, MZ, CS, ZRZ, JCH, EJR, SJL, DFW, LGH, LBN

Formal analysis: BJH, CS, ZRZ, JCH, LHW, PPT

Funding acquisition: PPT, JCH, MAH

Investigation: CRO, RWW, MZ, CS, ZRZ, JCH, EJR, SJL, DFW

Methodology: RWW, MZ, ZRZ, LHW, PPT, PC

Project administration: CRO, RWW, MZ, DJO, LGH, LBN, MAH

Resources: MAH

Supervision: BJH, CRO, RWW, MZ, DJO, RDS, MAH

Validation: BJH Visualization: BJH, LHW

Writing—original draft: BJH, RDS, MIB, MAH

Writing—review and editing: All authors

## Data Availability

Data may be made available for sharing through approach of the corresponding author and if permitted by the IRB.

## Acknowledgements

The authors gratefully acknowledge the essential contributions of faculty from the University of Iowa Roy J. and Lucille A. Carver College of Medicine Department of Orthopedics and Rehabilitation and Department of Neurosurgery who, in addition to the authors (CRO, RWW, MZ), participated in this study—Doctors Hitchon, Igram, Kawasaki, Oya, and Yamaguchi.

## Supplementary data

## Methods

### Biomarkers of Systemic Inflammation and Neuroinflammation

The lower limits of detection for each biomarker assay are summarized in Table S-1. Samples with biomarker concentrations calculated to be less than the lower limit of detection were used for analysis provided that the sample fluorescence signal was greater than the background fluorescence signal. In the two instances where the sample fluorescence signal was less than background, a default value of zero pg mL^-1^ was used.

**Table S1.** Plasma biomarker assay lower limits of detection.

| Biomarker | Lower Limit of Detection, $\text{pg mL}^{-1}$ |
| --- | --- |
| Interleukin-8 | 0.55 |
| Interleukin-10 | 2.60 |
| Tumor Necrosis Factor, alpha | 6.22 |
| Interleukin-6 | 0.63 |
| S100B | 150 |

## Results

### Blood loss and red blood cell transfusion

#### Blood loss

**Table S2.** Postoperative blood loss as a function of sex. Data are expressed as median (25^th^, 75^th^ percentiles)

| Sex | Postoperative blood loss per instrumented level, $\text{mL level}^{-1}$ | | *P* value | Effect Size (d) (95% CI) |
| --- | --- | --- | --- | --- |
|  | Tranexamic Acid | Placebo |  |  |
| Male | n=23: 125 (90, 178) | n=17: 200 (148, 268) | 0.050 | -0.68 (-1.30, -0.03) |
| Female | n=19: 147 (97, 189) | n=20: 223 (111, 254) | 0.136 | -0.61 (-1.20, 0.04) |
| Pooled | n=42: 128 (97, 186) | n=37: 207 (132, 258) | **0.013** | -0.64 (-1.09, -0.19) |

#### Red blood cell transfusion

Red blood cell (RBC) administration occurred in fewer participants who had instrumentation of ≤4 levels (TXA: 3/34=8.8%; Placebo: 7/37=18.9%; Combined: 10/71=14.1%) *vs* instrumentation of ≥5 levels (TXA: 7/9=78%; Placebo: 3/3=100%; Combined: 10/12=83%); *P*<0.001.

Among the 20 participants who received RBCs, 3/20 (15%) received RBCs intraoperatively only, 10/20 (50%) received RBCs both intra- and post-operatively, and 7/20 (35%) received RBCs postoperatively only. Postoperative RBC administration occurred more often in participants who had received intraoperative RBCs than in those who did not; (10/13=77%) *vs* (7/70=10%), respectively; *P*<0.000002. Thirty-two of 58 (55%) RBC units administered were administered postoperatively.

### Delirium

#### Delirium severity

Among 18 participants who had delirium, 9 (50%) had delirium onset on postoperative day 1. There was no association between the day of delirium onset and delirium onset severity score (Kendall’s τ= −0.221; *P*=0.274). Delirium severity score increased from 6 (4,7) at onset to a maximum of 9 (6, 12); *P*=0.0057. There was a positive association between delirium onset severity score and maximum delirium severity score (Kendall’s τ=0.5818; *P*=0.0015). Both maximum and average delirium severity scores were greater in participants who had delirium (n=18) compared to those who did not (n=47), both *P* <0.0000001.

**Table S3.** Delirium characteristics and outcomes. Data are expressed as median (25^th^, 75^th^ percentiles) or n (%)

| Variable | Delirium<br>(n = 18) | No Delirium<br>(n = 47) | *P*<br>value |
| --- | --- | --- | --- |
| Delirium onset: postoperative day n (%) |  |  |  |
| 1 | 9 (50%) |  |  |
| 2 | 4 (22%) |  |  |
| 3 | 5 (28%) |  |  |
| Delirium onset: severity score | 6 (4, 7) |  |  |
| Maximum delirium severity score | 9 (6, 12) | 1 (1, 2) | **<0.0000001** |
| Average delirium severity score | 4.2 (2.3, 5.6) | 1.0 (0.3, 1.2) | **<0.0000001** |
| Days with delirium, n (%) |  |  |  |
| 1 | 7 (39%) |  |  |
| 2 | 5 (28%) |  |  |
| 3 | 1 (6%) |  |  |
| 4 | 3 (17%) |  |  |
| 5 | 2 (11%) |  |  |
| Length of hospital stay, days | 7 (5, 8) | 3 (2, 4) | **0.000004** |
| Post-discharge, disposition, n (%) |  |  | **0.006** |
| Home | 8 (44%) | 39 (83%) |  |
| Inpatient rehabilitation | 9 (50%) | 7 (15%) |  |
| Skilled care | 1 (5.5%) | 1 (2.1%) |  |

Bold character highlights $P$ value <0.050.

Among the 18 participants who had delirium, 12 (66%) satisfied delirium diagnostic criteria for 1 or 2 days prior to discharge. There were positive associations between the total number of days with delirium and both delirium onset severity score (Kendall’s τ=0.4168; *P*=0.0323) and maximum delirium severity score (Kendall’s τ=0.7339; *P*=0.0001).

### Biomarkers of systemic inflammation and neuroinflammation

#### Cytokine/S100B interactions

In robust regression models, in the Placebo group (n=16), postoperative S100B concentrations were not associated with postoperative IL-10, TNFα, or IL-6 concentrations (all *P* values ≥0.163), with no significant cytokine*group interactions (all *P* values ≥0.188) indicating that TXA did not change the (absent) associations. In contrast, in the Placebo group, postoperative S100B concentrations were positively associated with postoperative IL-8 concentrations (*P*=0.047), with marginally negative IL-8*group interaction term (Beta = −45 [95% CI −95 to 3.9; *P*=0.073] suggesting TXA may have decreased the S100B response to IL8 (Fig. S1).

**Fig S1.**
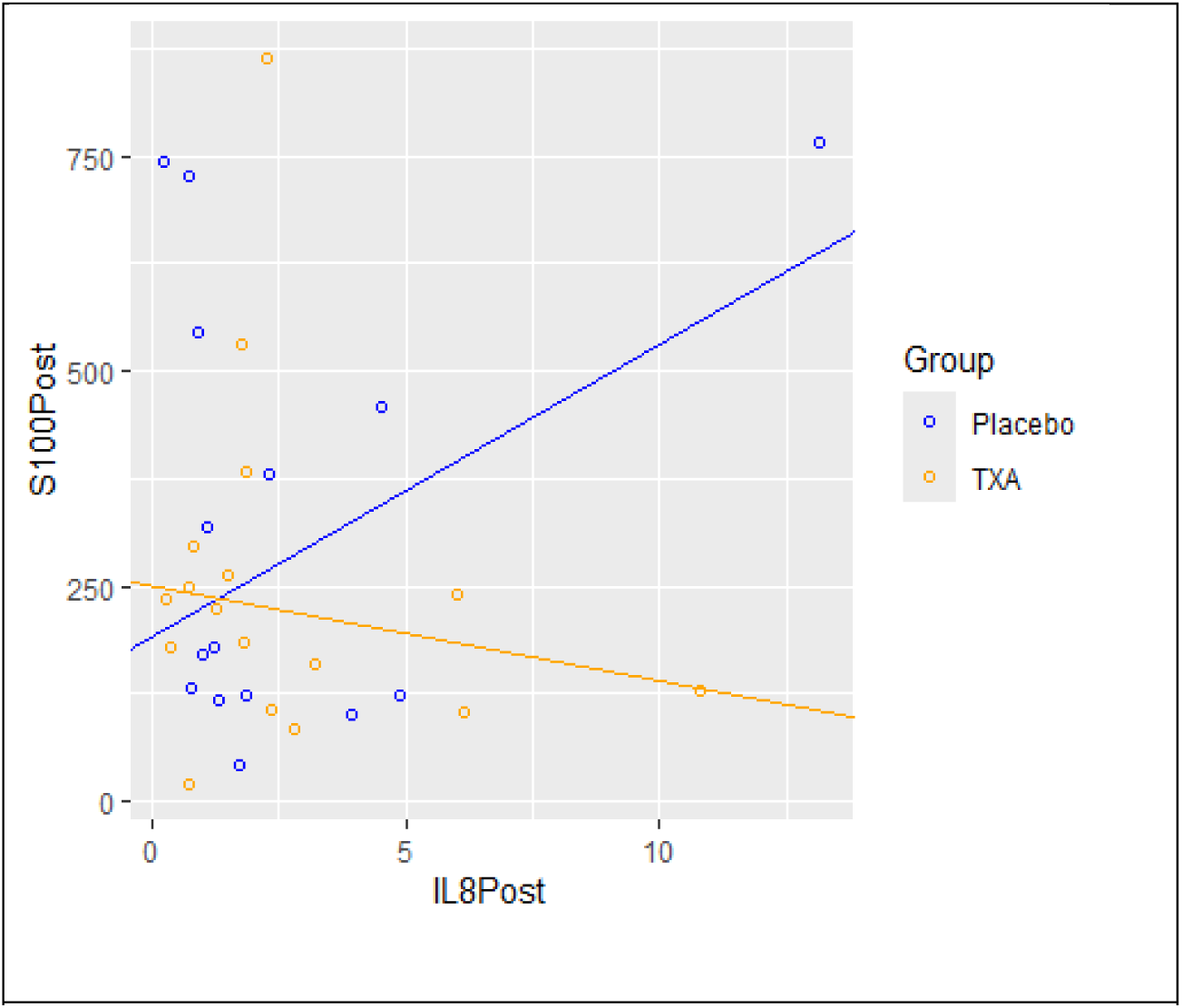
Postoperative S100B concentration *vs.* postoperative IL-8 concentration. TXA data points (n=16) are shown orange circles and lines and Placebo data points (n=16) Robust regression lines are shown.

### Cognitive Testing

Cognitive test scores of participants who did and did not have postoperative delirium are summarized in **Table S-4.** Among 18 participants who experienced postoperative delirium, there were no significant differences between preoperative and post-discharge test scores (all 4 *P* values ≥0.210). In contrast, in participants without delirium, there was a marginal but non-signficant post-discharge improvement in TMT-A scores (*P*=0.055) and significant improvements in post-discharge TMT-B (*P*=0.0004) and COWA (*P*=0.0006) scores.

**Table S4.** Cognitive test scores in participants with and without postoperative delirium. Data are expressed as median (25^th^, 75^th^ percentiles). * Because of small sample size (n=6), paired T-test.

| Cognitive Tests |  | Delirium | No Delirium |
| --- | --- | --- | --- |
| Telephone Interview of Cognitive Status-modified, score |  |  |  |
|  | n | 12 | 41 |
|  | Preoperative | 36 (30, 38) | 35 (33, 38) |
|  | Post-discharge | 35 (33, 40) | 36 (33, 40) |
|  | *P* value | 0.210 | 0.222 |
| Trail Making Test-Part A, z-score |  |  |  |
|  | n | 12 | 41 |
|  | Preoperative | -0.900 (-1.124, -0.374) | -0.964 (-1.382, -0.505) |
|  | Post-discharge | -0.823 (-1.011, -0.183) | -1.224 (-1.488, -0.622) |
|  | *P* value | 1 | 0.055 |
| Trail Making Test-Part B, z-score |  |  |  |
|  | n | 11 | 39 |
|  | Preoperative | -0.013 (-0.951, 0.858) | -0.109 (-0.931, 0.953) |
|  | Post-discharge | -0.059 (-0.354, 1.215) | -0.966 (-1.334, 0.161) |
|  | *P* value | 0.413 | **0.0004** |
| Controlled Oral Word Association, z-score |  |  |  |
|  | n | 6 | 30 |
|  | Preoperative | -0.551 (-0.874, -0.200) | -0.489 (-1.263, 0.736) |
|  | Post-discharge | -0.519 (-1.002, 0.290) | -0.187 (-0.800, 1.026) |
|  | *P* value | 0.922* | **0.0006** |

Bold character highlights *P* value <0.050.

## Footnotes

1 Exclusion of the two high value IL-6 outliers (one from each group) still resulted in a significant IL6*group interaction term; P=0.000007

